# Evaluation of a speech-based AI system for early detection of Alzheimer’s disease remotely via smartphones

**DOI:** 10.1101/2021.10.19.21264878

**Authors:** Emil Fristed, Caroline Skirrow, Marton Meszaros, Raphael Lenain, Udeepa Meepegama, Stefano Cappa, Dag Aarsland, Jack Weston

## Abstract

**Background:** Changes in speech, language, and episodic and semantic memory are documented in Alzheimer’s disease (AD) years before routine diagnosis.

**Aims:** Develop an Artificial Intelligence (AI) system detecting amyloid-confirmed prodromal and preclinical AD from speech collected remotely via participants’ smartphones.

**Method:** A convenience sample of 133 participants with established amyloid beta and clinical diagnostic status (66 A***β***+, 67 A***β***-; 71 cognitively unimpaired (CU), 62 with mild cognitive impairment (MCI) or mild AD) completed clinical assessments for the AMYPRED study (NCT04828122). Participants completed optional remote assessments daily for 7-8 days, including the Automatic Story Recall Task (ASRT), a story recall paradigm with short and long variants, and immediate and delayed recall phases. Vector-based representations from each story source and transcribed retelling were produced using ParaBLEU, a paraphrase evaluation model. Representations were fed into logistic regression models trained with tournament leave-pair-out cross-validation analysis, predicting A***β*** status and MCI/mild AD within the full sample and A***β*** status in clinical diagnostic subsamples.

**Findings:** At least one full remote ASRT assessment was completed by 115 participants (mean age=69.6 (range 54-80); 63 female/52 male; 66 CU and 49 MCI/mild AD, 56 A***β***+ and 59 A***β***-). Using an average of 2.7 minutes of automatically transcribed speech from immediate recall of short stories, the AI system predicted MCI/mild AD in the full sample (AUC=0.85 +/- 0.08), and amyloid in MCI/mild AD (AUC=0.73 +/- 0.14) and CU subsamples (AUC=0.71 +/- 0.13). Amyloid classification within the full sample was no better than chance (AUC=0.57 +/- 0.11). Broadly similar results were reported for manually transcribed data, long ASRTs and delayed recall.

**Interpretation:** Combined with advanced AI language models, brief, remote speech-based testing offers simple, accessible and cost-effective screening for early stage AD.

**Funding:** Novoic.

**Research in context:** *Evidence before this study:* Recent systematic reviews have examined the use of speech data to detect vocal and linguistic changes taking place in Alzheimer’s dementia. Most of this research has been completed in the DementiaBank cohort, where subjects are usually in the (more progressed) dementia stages and without biomarker confirmation of Alzheimer’s disease (AD). Whether speech assessment can be used in a biomarker-confirmed, early stage (preclinical and prodromal) AD population has not yet been tested. Most prior work has relied on extracting manually defined “features”, e.g. the noun rate, which has too low a predictive value to offer clinical utility in an early stage AD population. In recent years, audio- and text-based machine learning models have improved significantly and a few studies have used such models in the context of classifying AD dementia. These approaches could offer greater sensitivity but it remains to be seen how well they work in a biomarker-confirmed, early stage AD population. Most studies have relied on controlled research settings and on manually transcribing speech before analysis, both of which limit broader applicability and use in clinical practice.

*Added value of this study:* This study tests the feasibility of advanced speech analysis for clinical testing of early stage AD. We present the results from a cross-sectional sample in the UK examining the predictive ability of fully automated speech-based testing in biomarker-confirmed early stage Alzheimer’s disease. We use a novel artificial intelligence (AI) system, which delivers sensitive indicators of AD-at-risk or subtle cognitive impairment. The AI system differentiates amyloid beta positive and amyloid beta negative subjects, and subjects with mild cognitive impairment (MCI) or mild AD from cognitively healthy subjects. Importantly the system is fully remote and self-contained: participants’ own devices are used for test administration and speech capture. Transcription and analyses are automated, with limited signal loss. Overall the results support the real-world applicability of speech-based assessment to detect early stage Alzheimer’s disease. While a number of medical devices have recently been approved using image-based AI algorithms, the present research is the first to demonstrate the use case and promise of speech-based AI systems for clinical practice.

*Implications of all the available evidence:* Prior research has shown compelling evidence of speech- and language-based changes occurring in more progressed stages of Alzheimer’s disease. Our study builds on this early work to show the clinical utility and feasibility of speech-based AI systems for the detection of Alzheimer’s disease in its earliest stages. Our work, using advanced AI systems, shows sensitivity to a biomarker-confirmed early stage AD population. Speech data can be collected with self-administered assessments completed in a real world setting, and analysed automatically. With the first treatment for AD entering the market, there is an urgent need for scalable, affordable, convenient and accessible testing to screen at-risk subject candidates for biomarker assessment and early cognitive impairment. Sensitive speech-based biomarkers may help to fulfil this unmet need.

## Introduction

Pathological changes in Alzheimer’s disease (AD) begin years before symptoms of dementia or the early clinical stages of mild cognitive impairment (MCI), and up to decades before diagnosis^1^. Clinical trials targeting the earliest stages of AD typically rely on measuring Amyloid beta (A***β***) biomarkers using positron emission tomography (PET) or in cerebrospinal fluid (CSF) obtained from lumbar puncture. The high cost and/or invasive nature of these procedures restricts use in standard clinical care and broader population screening. Blood plasma biomarkers hold promise for reducing screening costs but remain invasive and do not differentiate clinical stages of the disease^2^.

More importantly, cognitively unimpaired individuals with biomarker evidence of both amyloid beta and tau pathology will not always develop clinical manifestations in their lifetime, and should only be considered at-risk for progression to AD, reserving diagnosis of AD to people with evidence of an AD cognitive phenotype^3^. Cognitive testing is thus crucial for an early diagnosis. Cognitive tests have been supported for use as endpoints of treatment efficacy early in the Alzheimer’s continuum by regulatory bodies^4,5^. However, traditional cognitive tests typically require significant qualified staff time to administer and score. In the case of amyloid positive asymptomatic subjects, only subtle impairments or longitudinal change are observable^6,7^.

Cognitive test results often reflect simple indices of response accuracy or recall, ignoring differences in the content, structure and delivery of patients’ responses to tasks. For episodic memory tests, such as tests of story recall, test performance does not typically differ between clinically unimpaired A***β***+ and A***β***-individuals, but differences can be seen in the recall of proper nouns^8^, and the serial position of elements recalled^9^. Later in the disease course, differences are seen in rates of verbatim or paraphrased recall^10^, language density and pauses^11^.

There is a need for cognitive screening tools allowing fast and frequent assessment of the at-risk population. Speech data collected on ubiquitous digital devices represents an excellent candidate for this goal. Verbal memory tasks can be scored automatically using natural language processing technologies^12^, and augmented with acoustic and linguistic measures to further improve detection^11^. Recent methods in artificial intelligence (AI) enable extraction of more information-dense patterns from text data^13,14^. We hypothesise that these could form the basis of speech biomarkers sensitive to earlier disease stages, possibly before overt cognitive decline (asymptomatic at-risk individuals).

Using speech elicited from a remotely self-administered story recall task, we aim to develop an AI-based system to (1) differentiate A***β***+ and A***β***-subjects; (2) differentiate those with and without mild cognitive impairment. The test would be usable remotely for initial clinical screening to detect MCI and subtle signs of cognitive decline in amyloid-confirmed asymptomatic subjects (preclinical AD). Furthermore, we examine the performance of the index test compared to current standard-of-care in primary care referrals for MCI using a simulation approach.

## Methods

### Study Design

The AMYPRED study (NCT04828122) is a prospective study with data collection planned before the index test was performed. The study uses a 2×2 cross-sectional design, combining amyloid status (A***β***+ and A***β***-) and clinical status (cognitively unimpaired (CU) and MCI/mild AD). Reference standards for A***β*** positivity and clinical status were established prior to recruitment into the study.

### Participants

Participants were a convenience sample recruited from trial participant registries in three UK sites (London/Guildford, Plymouth and Birmingham) between November 2020 to July 2021. Subjects were approached if they had undergone a prior A***β*** PET scan or CSF test (confirmed A***β***-within 30 months or A***β***+ within 60 months) and were cognitively unimpaired (CU) or diagnosed with MCI or mild AD in the previous 5 years. MCI due to AD and mild AD diagnoses were made following National Institute of Aging-Alzheimer’s Association core clinical criteria^15^.

Potential participants were screened via video conferencing, during which the Mini-Mental State Exam (MMSE)^16^ was administered. Inclusion criteria comprised: age 50-85; MMSE score 23-30 for participants with MCI/mild AD, 26-30 for CU; clinical diagnosis made in previous 5 years for participants with MCI/mild AD; English as a first language; availability of a caregiver or close associate to support completing the Clinical Dementia Rating scale (CDR) semi-structured interview^17^; ability to use and access to a smartphone (Android 7 or above or iOS 11 or above); and access to the internet on a personal computer, notebook or tablet (supported operating systems and internet browser software documented in supplementary materials).

Exclusions comprised: current diagnosis of general anxiety disorder or major depressive disorder; recent (6-month) history of unstable psychiatric illness; history of stroke within the past 2 years or transient ischaemic attack or unexplained loss of consciousness in the last 12 months. Participants taking medications for AD symptoms were required to be on a stable dose for at least 8 weeks.

### Study assessments

Participants underwent a clinical assessment via video call, followed by optional remote assessments daily using their personal digital devices for 7-8 days.

#### Telemedicine assessments

Cognitive tests part of the Preclinical Alzheimer’s Cognitive Composite with semantic processing (PACC5) were administered and mean z-score was calculated as previously described^6^. The CDR^17^, assessing the severity of cognitive symptoms of dementia, was completed by experienced research staff and scored to deliver the CDR Global Score (CDR-G). Task modifications enabling remote assessment during the SARS-CoV-2 pandemic are detailed in supplementary materials.

#### Remote assessments

During telemedicine assessments, participants were supported to install the Novoic mobile application on their own smartphone. They were encouraged to complete optional unsupervised self-assessments daily for the following 7-8 days. Remote self-assessments included Automatic Story Recall Tests (ASRT), consisting of 18 short and 18 long story variants (mean of 119 and 224 words per story, and stimulus duration approx 1 minute and 1min 40 seconds, respectively). ASRTs were administered in triplets (three stories administered consecutively each day). The self-assessment schedule is provided in supplementary materials.

Participants listened to pre-recorded ASRTs and retold stories in as much detail as they could remember, immediately after presentation of each story and after a delay. Task responses were recorded on the app and automatically uploaded to a secure server.

### Sample size determination

Minimal bounds were set on the dataset size required to train flexible models with many parameters but strong representations (20 participants per group). Power calculations completed using the pROC package in R, with power specified at 80% and significance set at 0.05, indicated that for full sample analyses an AUC of 0.67 would be detectable with n=40 participants in each group. Using similar parameters, an AUC of 0.74 would be detectable with a n=20 participants in smaller subgroup analyses.

### Outcome measures

Key outcome measures included the AI-based index test result, identifying: (1) A***β*** positivity in the full sample; (2) MCI in the full sample; (3) A***β*** positivity in MCI/mild AD; (4) A***β*** positivity in the CU subsample. Furthermore, an AI-based continuous measure predicting PACC5 scores was derived. Diagnostic accuracy was established through comparison with PET or CSF A***β*** status and clinical diagnosis established in prior trials.

Short ASRT triplets (immediate recall, automatically transcribed) were primary measures of interest. These were experienced as lower burden by participants, yielding higher compliance and a greater number of datapoints for model training and analysis. Long ASRT stories and delayed recall were also examined.

### Oversight

This study was approved by Institutional Review at the West Midlands Health Research Authority (UK REC reference: 20/WM/0116). Informed consent was taken electronically in accordance with HRA guidelines.

### Overview of the AI system

The AI system was based on the “edit encoder” of the ParaBLEU model^18^, the state-of-the-art for paraphrase evaluation. Given two input texts, the edit encoder outputs a vector-based representation of the abstract, generalized patterns that differ between them. On an established paraphrase quality benchmark, models using ParaBLEU numerical representations correlate more strongly with human judgements than other existing metrics^18^. Differing from the standard ParaBLEU setup, the model was pretrained with longer paraphrase examples to mirror the length of source-retelling pairs, and without the entailment component of the loss function as entailment labels were unavailable for the updated pretraining dataset. The base model of the edit encoder used a pretrained Longformer model rather than a pretrained RoBERTa model, to accommodate longer texts.

### Statistical analysis

#### AI system application

Although adherence varied across participants, ASRTs have high parallel forms reliability, and modest practice effects, indicating that story triplets can be substituted for one another (Skirrow et al., *submitted*)^19^. One complete long and one complete short triplet (comprising six retellings per triplet: three immediate and three delayed) were picked randomly and uniformly for each participant across assessment days for onward analysis. Long and short triplets and immediate and delayed recall were examined separately.

Responses were transcribed manually and with an out-of-the-box automatic speech recognition (ASR) system. Analyses were completed in Python, using a proprietary framework built using PyTorch.

For each retelling, two representations were derived, based on non-redundant differences between the target (story text) and retelling (target→retelling and retelling→target) as represented by the ParaBLEU model. Classifiers were trained using logistic regression models with the sklearn package in Python to predict pairs of labels (MCI/mild AD or CU; A***β***+ or A***β***-) with tournament leave-pair-out cross-validation analysis (TLPO)^20^. The TLPO process was run twice - once per representation - to obtain two sets of participant-level scores. These are ensembled by simple averaging. To augment the training sample, participants completing any remote assessments (including only partially completed ASRT triplets) were included in model training. Predictions were tested only on participants with complete data.

### Clinical and biomarker discrimination of models

Participant-level scores were ensembled across the three stories per triplet (immediate or delayed recall) to create participant level predictions. These were used to create a ranking for receiver operating characteristic (ROC) curve analysis. The AI system was compared to two comparison models, (1) a demographic comparison (age, sex and years of education) and (2) the PACC5 z-score. For n=1 CU participant, missing data for years in education was replaced with the group median. Comparison models were trained using the participant information as input(s) to a logistic regression model using an identical setup to the models trained on top of the ParaBLEU representations. Predictions were assessed by the area under the curve (AUC); and sensitivity, specificity and Cohen’s kappa at Youden’s index for the test result in comparison with reference standards. Statistical significance of differences between AUCs and 95% confidence intervals for AUCs were computed using DeLong’s method^21^.

### PACC5 prediction

PACC5 z-scores were predicted from speech samples, trained with leave-one-out cross-validation using ridge regression models with polynomial kernels. The Pearson correlation coefficient between predicted and actual PACC5 scores was computed.

### Screening simulation

Screening for MCI was simulated in a hypothetical age 65+ sample (n=1000) with proportional representation of each age group representative of the US population^22^, and MCI prevalence estimates by age from prior meta-analysis^23^. The AI system’s (short stories, immediate recall, ASR) sensitivity and specificity within the sample was determined at Youden’s index, and compared to physician subjective judgement and MMSE reported in prior research^24^. Methods are described in supplementary materials.

### Role of funding source

The study was funded by Novoic, a clinical late-stage digital medtech company developing AI-based speech biomarkers. The funder of the study provided financial support towards collection and analysis of the data and was involved in study design, data interpretation and writing of the report.

## Results

Participant recruitment is presented in figure 1. One hundred and thirty-three participants were recruited and completed study visits via video call, with 86% with A***β*** status confirmed by PET scan (115/133). The MCI/mild AD participant group comprised primarily MCI participants, with ten individuals (20.4%) having a diagnosis of mild AD.

**Figure 1:**
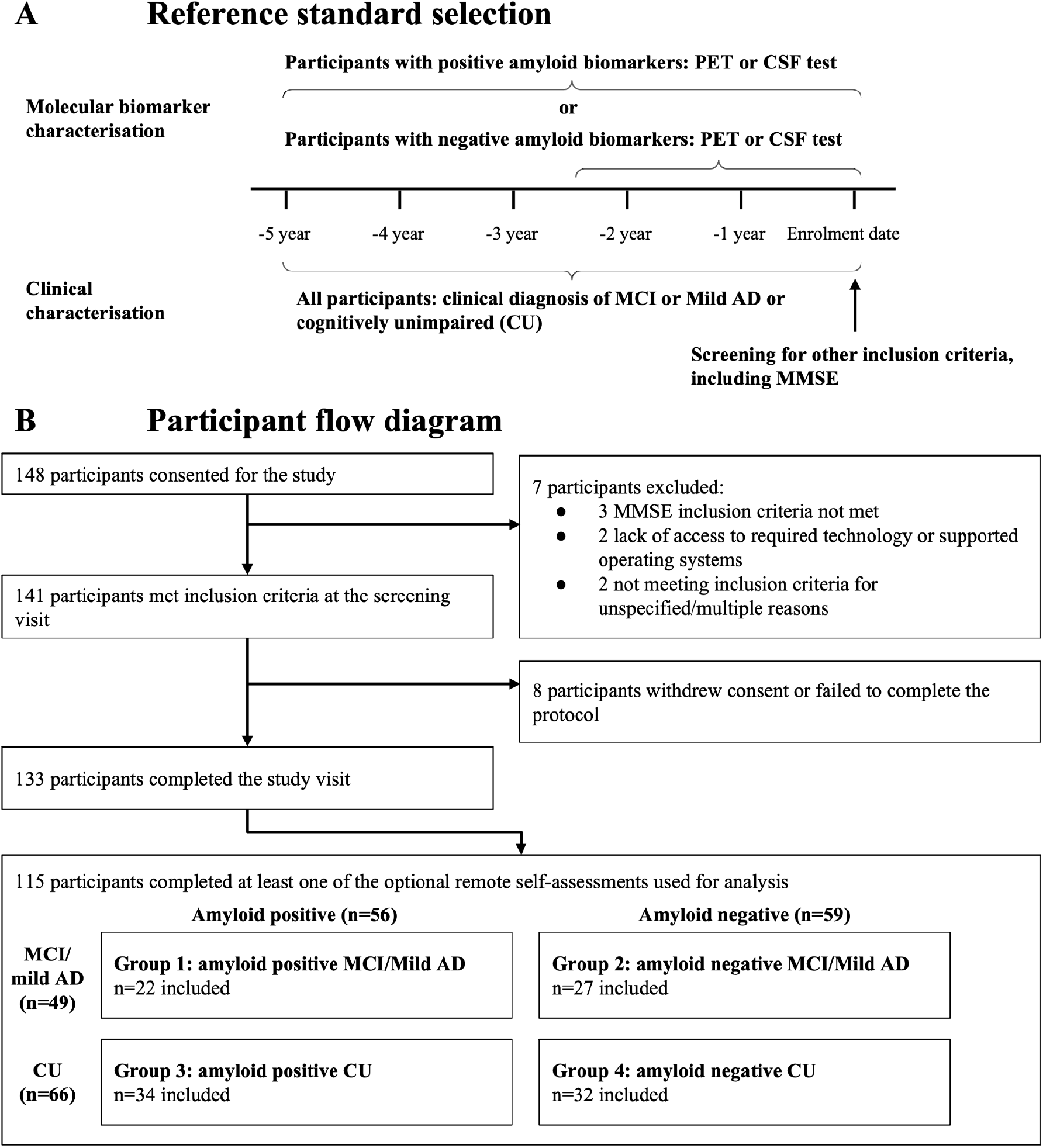
Patient and reference standard selection. (A) Participant inclusion criteria: participants were included based on prior amyloid status and clinical diagnosis confirmation before being screened for the study’s other inclusion criteria. (B) Participant flow diagram, documenting exclusions and dropouts during study recruitment. PET: positron emission tomography; CSF: cerebrospinal fluid; CU: cognitively unimpaired; MCI: mild cognitive impairment; AD: Alzheimer’s disease; MMSE: Mini-Mental State Exam

At least one full optional remote self-assessment was completed by 86% (115/133). For those who engaged in at least one full optional remote self-assessment, overall engagement with daily testing was high (mean of 77% in full sample; 78% in CU and 66% in MCI/mild AD).

Those who did not complete remote assessments were more commonly diagnosed with MCI/mild AD (χ^2^=5.49, p=0.01) and had lower CDR-G scores (r=-0.19, p=0.04). However, they did not differ in age (r=-0.15, p-0.12), education level (r=-0.005, p=0.96), male/female ratio (χ^2^=0.004, p=0.95), A***β***+/A***β***-ratio (χ^2^=0.96, p=0.33) or MMSE score (r=-0.15, p=0.11).

Demographics in the remote assessment sample (for subgroup and full sample analyses) are shown in table 1. This shows no clear differences between research, clinical and biomarker groups. Demographics for the entire sample and by short and long ASRT training sets are given in supplementary tables s3-s5.

**Table 1:** Participant demographic and clinical characteristics. Demographic and clinical characteristics shown by research groupings 1-4, and summary statistics for participants characterised by clinical diagnostic or biomarker profiles. MCI: mild cognitive impairment; AD: Alzheimer’s dementia; CU: cognitively unimpaired; N, number; SD, standard deviation. Group 1: amyloid beta positive MCI/mild AD, Group 2: amyloid beta negative MCI/mild AD, Group 3: amyloid beta positive cognitively unimpaired; Group 4: amyloid beta negative cognitively unimpaired; MMSE: Mini-Mental State Exam; CDR-G: Clinical Dementia Rating Scale - Global Score.

|  | Subgroup analyses |  |  |  |  | Full sample analyses |  |  |  |  |  |
| --- | --- | --- | --- | --- | --- | --- | --- | --- | --- | --- | --- |
|  | Group 1:<br>(N=22) | Group 2:<br>(N=27) | Group 3:<br>(N=34) | Group 4:<br>(N=32) | p-value | Clinical group |  |  | Biomarker group |  |  |
|  |  |  |  |  |  | CU<br>(N=66) | MCI/mild<br>(AD<br>N=49) | p-value | Amyloid<br>beta<br>negative<br>(N=59) | Amyloid<br>beta<br>positive<br>(N=56) | p-value |
| Amyloid beta<br>positive/<br>negative (N) | Positive | Negative | Positive | Negative | - | 34/32 | 22/27 | 0.48 | Negative | Positive | - |
| MCI/CU<br>group (N) | MCI/Mild<br>AD | MCI/Mild<br>AD | CU | CU | - | CU | MCI | - | 27/32 | 22/34 | 0.48 |
| Female/male<br>(N) | 7/15 | 16/11 | 21/13 | 19/13 | 0.12 | 40/26 | 23/26 | 0.15 | 35/24 | 28/28 | 0.32 |
| Years in<br>education,<br>mean (SD) | 15.05<br>(3.32) | 15.08<br>(2.92) | 14.97<br>(3.77) | 15.41<br>(3.35) | 0.98 | 15.18<br>(3.55) | 15.06<br>(3.08) | 0.78 | 15.26<br>(3.14) | 15.00<br>(3.57) | 0.78 |
| Age, mean<br>(SD) | 71.00<br>(5.83) | 67.22<br>(7.95) | 70.44<br>(4.18) | 69.84<br>(3.78) | 0.38 | 70.15<br>(3.97) | 68.92<br>(7.26) | 0.74 | 68.64<br>(6.14) | 70.66<br>(4.85) | 0.13 |
| MMSE, mean<br>(SD) | 27.27<sup>A</sup><br>(1.64) | 27.41<sup>B</sup><br>(2.02) | 29.24<sup>C</sup><br>(1.05) | 28.81<sup>D</sup><br>(1.11) | <0.001<br>AC, AD, BC,<br>BD | 29.03<br>(1.09) | 27.33<br>(1.85) | <0.001 | 28.15<br>(1.73) | 28.46<br>(1.62) | 0.25 |
| CDR-G,<br>mean (SD) | 0.52<sup>A</sup><br>(0.11) | 0.50<sup>B</sup><br>(0.14) | 0.08<sup>C</sup><br>(0.18) | 0.10<sup>D</sup><br>(0.20) | <0.001<br>AC, AD, BC,<br>BD | 0.09<br>(0.19) | 0.51<br>(0.12) | <0.001 | 0.28<br>(0.27) | 0.25<br>(0.27) | 0.54 |

### Outcome measures

#### AI system application

A***β*** classification in the full sample was no better than chance across the AI system and the two comparison analyses (figure 2A). Within the MCI (figure 2C) and CU (figure 2D) subsamples the AI system AUC for A***β*** detection was 0.73, and 0.71 respectively, showing better A***β*** signal when examined within more homogeneous groups. MCI classification using the AI system in the full sample yielded an AUC of 0.85 (figure 2B).

**Figure 2:**
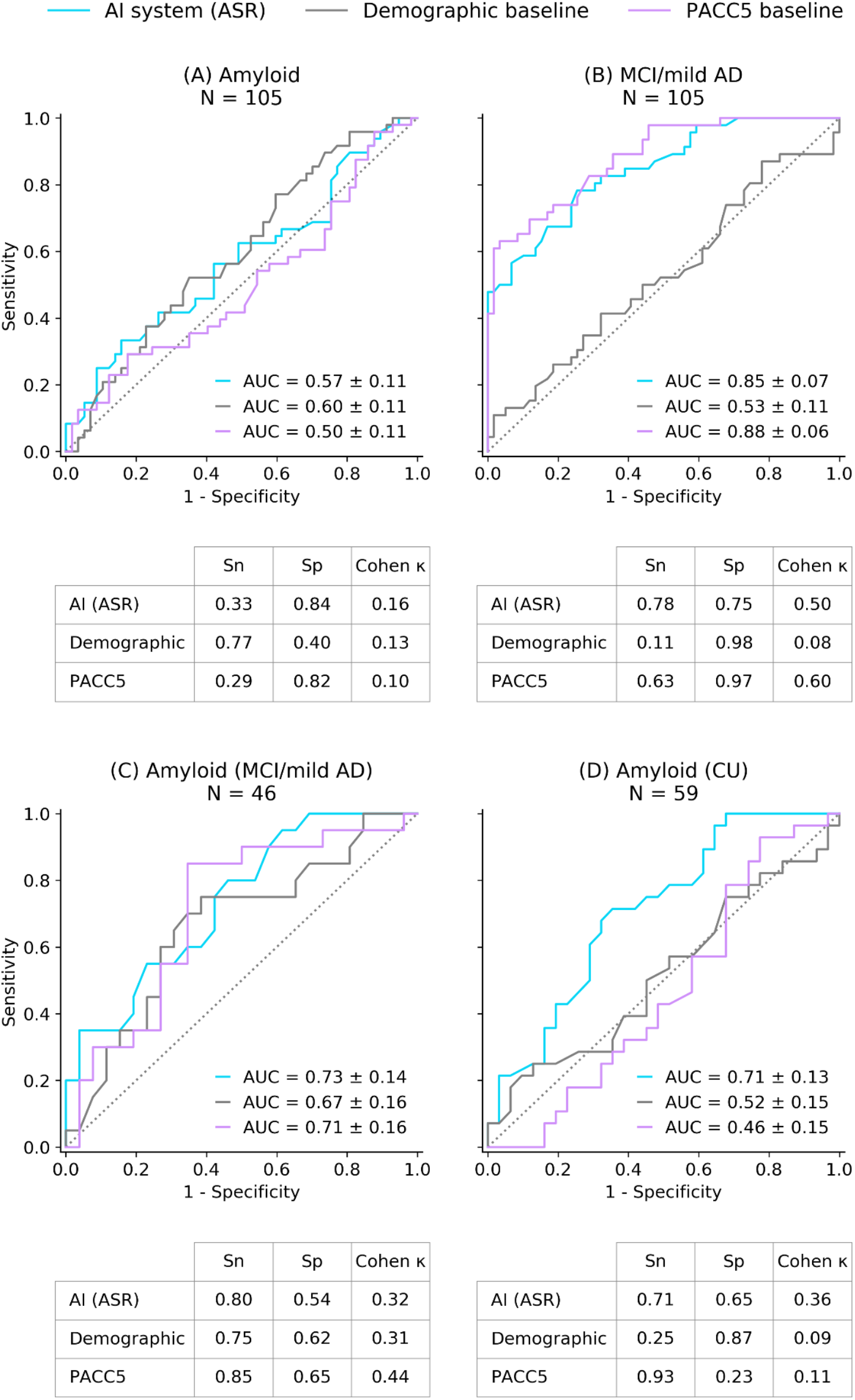
ROC curves for the AI system and comparison models (short ASRTs, immediate recall) AUCs for the classifiers predicting: (A) amyloid, (B) MCI/mild AD in the full sample. Subsample comparisons of classifier performance predicting (C) amyloid within the MCI/mild AD; and (D) amyloid in the cognitively unimpaired (CU) sample. The table below each figure provides sensitivity and specificity at Youden’s index and Cohen’s kappa measures. The reference test was biomarker confirmation on PET or CSF for A, C and D. Reference test was clinical diagnosis MMSE inclusion criteria for B. The demographic comparison includes age, sex and education level. ASR: automatic speech recognition - automatically transcribed; MCI: mild cognitive impairment; CU: cognitively unimpaired; AI: artificial intelligence; ASRT: automatic story recall test; PACC5: preclinical Alzheimer’s cognitive composite with semantic processing; ROC: receiver operator characteristic (curve); AUC: area under the curve.

AI system performance did not differ depending on automatic versus manual transcription, and results were broadly consistent for long ASRT stories and delayed recall (table 2; supplementary figures). Demographic comparison performed consistently worse than the AI system (figure 2, table 2), with confidence intervals incorporating chance level, with the exception of amyloid prediction in the MCI subsample. As compared with the AI system, the PACC5 comparison delivered subtly but non-significantly higher AUCs for detecting MCI in the full sample, but similar performance for A***β*** status in MCI and poorer performance for A***β*** status in CU (figure 2).

**Table 2:** Area under the curve (AUC +/- 95% confidence intervals) for ASRT variants and demographic comparison. Comparison of performance of the AI system for classifying (A) Amyloid beta in the full sample, (B) MCI in the full sample, (C) Amyloid beta in the MCI subsample, and (D) Amyloid beta in the CU subsample, using immediate and delayed recalls of short and long ASRT triplets as input. Difference between AI system and demographic comparison *p<0.0001. CU: cognitively unimpaired; MCI: mild cognitive impairment; ASRT: automatic story recall task; ASR: automatic speech recognition - automatically transcribed; manual: manually transcribed.

| Task type | Delay type | Model | (A) Full sample amyloid beta | (B) Full sample MCI | (C) Amyloid beta in MCI subsample | (D) Amyloid beta in CU group |
| --- | --- | --- | --- | --- | --- | --- |
| Short ASRT | Immediate | Sample size | n = 105 | n = 105 | n = 46 | n = 59 |
|  |  | AI system (ASR) | 0.57 +/- 0.11 | 0.85 +/- 0.07* | 0.73 +/- 0.14 | 0.71 +/- 0.13 |
|  |  | AI system (manual) | 0.56 +/- 0.12 | 0.83 +/- 0.08* | 0.74 +/- 0.14 | 0.69 +/- 0.13 |
|  |  | Demographic comparison | 0.60 +/- 0.11 | 0.53 +/- 0.11 | 0.67 +/- 0.16 | 0.52 +/- 0.15 |
| Short ASRT | Delayed | Sample size | n = 105 | n = 105 | n = 46 | n = 59 |
|  |  | AI system (ASR) | 0.59 +/- 0.11 | 0.84 +/- 0.08* | 0.68 +/- 0.16 | 0.70 +/- 0.13 |
|  |  | AI system (manual) | 0.61 +/- 0.11 | 0.83 +/- 0.08* | 0.69 +/- 0.16 | 0.71 +/- 0.14 |
|  |  | Demographic comparison | 0.60 +/- 0.11 | 0.53 +/- 0.11 | 0.67 +/- 0.16 | 0.52 +/- 0.15 |
| Long ASRT | Immediate | Sample size | n = 97 | n = 97 | n = 39 | n = 58 |
|  |  | AI system (ASR) | 0.59 +/- 0.12 | 0.85 +/- 0.08* | 0.85 +/- 0.13 | 0.53 +/- 0.15 |
|  |  | AI system (manual) | 0.61 +/- 0.12 | 0.87 +/- 0.07* | 0.91 +/- 0.09 | 0.59 +/- 0.15 |
|  |  | Demographic comparison | 0.62 +/- 0.11 | 0.57 +/- 0.12 | 0.75 +/- 0.16 | 0.47 +/- 0.15 |
| Long ASRT | Delayed | Sample size | n = 97 | n = 97 | n = 39 | n = 58 |
|  |  | AI system (ASR) | 0.55 +/- 0.12 | 0.83 +/- 0.08* | 0.86 +/- 0.12 | 0.53 +/- 0.15 |
|  |  | AI system (manual) | 0.59 +/- 0.12 | 0.85 +/- 0.08* | 0.88 +/- 0.10 | 0.50 +/- 0.15 |
|  |  | Demographic comparison | 0.62 +/- 0.11 | 0.57 +/- 0.12 | 0.75 +/- 0.16 | 0.45 +/- 0.15 |

#### PACC5 prediction

The Pearson correlation between AI-model predicted and actual PACC-5 z scores was 0.74.

#### Simulation of MCI screening in primary care

In a simulated population sample age 65+ (MCI prevalence 16%), in comparison with unassisted physician judgement^24^, routine screening using the AI system (short stories, immediate recall) would increase correct referral rates in primary care by 56.0%, and reduce incorrect referrals by 26.5%; in comparison with screening via the MMSE^24^ the AI system would increase correct referral from primary care by 52.9%.

## Discussion

Research documents changes in vocal and linguistic speech patterns in AD, primarily in cohorts with more progressed AD and without biomarker confirmation^13,25^. The current findings show changes in speech occurring earlier in the disease process. Furthermore, we find modest but detectable differences in speech relating to changes associated with A***β*** positivity.

For the lowest burden assessments (short stories, immediate recall), the AI system predicted MCI (AUC 0.85) and A***β*** positivity in MCI and CU participant groups (AUC 0.73 and 0.71, respectively), but with A***β*** predictions in the full sample being no better than random. This could be due to more subtle impairments associated with A***β*** positivity, which may be obscured by broader changes seen accompanying MCI. The AI system consistently outperformed the demographic comparison and performed as well as, or better than, a lengthy supervised test battery developed to detect cognitive changes in preclinical AD (PACC5).

AI system results were consistent across manual and ASR transcription. MCI prediction was consistent for long and short ASRTs and immediate and delayed recall. A***β*** status prediction was not as consistent across task variants and groups: in cognitively unimpaired participants the AI system performed well for short, but not long, ASRTs; predictions of A***β*** status in MCI was above random for all task variants, but in the presence of an elevated demographic comparison. These results may indicate differential task difficulty effects interacting with demographic and clinical-biomarker groupings.

In the context of potential improvement in outcomes through lifestyle and medical interventions^26^, and the availability of new amyloid-targeting medications, early detection of AD and clear disease indication matters. However, in clinical practice, AD is not routinely screened for^27^ and is underdiagnosed even at the dementia stage^28^. Compared with standard-of-care assessments for MCI, routine screening using the AI system could increase correct referrals by up to 56.0% and reduce incorrect referrals by 26.5%.

Speech assessments were unsupervised, self-administered and analysed with an automated pipeline. Remote, unsupervised testing can improve inclusivity, increase standardisation and provide access to more advanced testing without the need for extensive experience of neuropsychological workup. Furthermore, speech-based AI models present as a potentially attractive low-cost and low-burden screen for A***β*** positivity. Combining the algorithm with other risk factors (e.g. age, APOE genotype) could further increase discriminative power.

### Limitations

We recruited participants with prior amyloid PET and CSF amyloid test results and clinical diagnoses. With increasing A***β*** positivity with age^29^, conversion may have occurred for some participants in the interim period. CSF and PET A***β*** positivity are differentially associated with cognitive decline, suggesting that they may be optimally sensitive at different disease stages^30^. Similarly, variation in diagnostic criteria for MCI/Mild AD (between trials where participants were recruited from) is likely to have introduced variability in our diagnostic reference standards. Even a small number of false labels can impact training of AI systems, and improvements in model performance could be expected with concurrent and consistent reference standards.

Although uptake of optional remote assessment was high, non-completion was associated with greater CDR-G indexed clinical impairment and was more common in MCI/Mild AD participants. Remote, unsupervised cognitive assessments may be challenging for individuals with more progressed cognitive impairment. For these subjects, supervised testing in clinic or via telemedicine may be more appropriate.

Test engagement varied from day to day and, as a result, our analyses included test results from different ASRT stimuli and testing days across different participants. Variability introduced by differences in the story stimuli themselves and practice effects, may have affected sensitivity of the AI system. However, this approach allowed us to maximise the sample available, and enabled us to develop stimulus-agnostic AI models, which can be applied across a class of stimuli.

Our AI system was developed and tested within a British English speaking sample, selected to exclude concurrent neurological and mental health conditions. Validation is now needed in more clinically heterogeneous samples and across different accents and languages. Larger-scale studies are needed to confirm and refine our results. We expect significant performance improvements in the AI system with a larger training dataset, increasing power to detect more subtle changes in speech patterns.

## Conflict of interest statement

EF, JW, MM, CS, RL and UM are employees of Novoic Ltd. EF, JW and MM are shareholders and MM, RL, UM, and CS are option holders in the company. EF and JW are directors on the board of Novoic. SC has received speaker’s fees from Roche and Biogen. DA has received research support and/or honoraria from Astra-Zeneca, Lundbeck, Novartis Pharmaceuticals, Evonik, Roche Diagnostics, and GE Health, and served as paid consultant for H. Lundbeck, Eisai, Heptares, Mentis Cura, Eli Lilly, Cognetivity, Enterin, Acadia, and Biogen.

## Author contributions

The study protocol was designed by EF, MM, and JW. The research study and data collection was coordinated by MM. Software was developed by JW, UM, RL and EF. Underlying data was verified by all authors (data acquisition and completeness MM, database management UM, quality control CS, data generated by analytic models JW, EF, UM, and RL). Analyses were completed by RL, CS, UM, JW, and EF. The first draft of the manuscript was completed by EF and CS. All authors contributed to the revision of the manuscript.

## Data sharing

Speech data is identifiable and cannot be shared, but all deidentified quantitative participant data shown in the present study are available upon reasonable request to the authors.

## Notes

### Clinical Trial

NCT04828122

### Author Declarations

Ethics committee of the West Midlands Health Research Authority gave ethical approval for this work.

### Summary of Updates

Reporting error on recruitment statistics has been corrected.

