## supplementary for "Evaluation of a speech-based AI system for early detection of Alzheimer’s disease remotely via smartphones"

### Supplementary material:

#### 1. Supported operating systems

Supported operating systems and internet browser software:

- macOS X with macOS 10.9 or later; or
- Windows 7 or above; or
- Ubuntu 12.04 or higher; or
- Have access to one of following internet browser software:
    - Internet Explorer version 11 or above; or
    - Microsoft Edge version 12 or above; or
    - Firefox version 27 or above; or
    - Google Chrome version 30 or above; or
    - Safari version 7 or above.
- Capable of audio and video recording.
- Able to connect to the internet.

#### 2. Telemedicine assessments

All participants were assessed via telemedicine to minimise participant risk during the context of the SARS-Cov-2 pandemic, which was ongoing during the study. GDPR-compliant Zoom video conferencing software was used, due to its robust security features.

Tests required for the Preclinical Alzheimer's Cognitive Composite with semantic processing (PACC5) were administered and mean z-score was calculated as previously described to examine early AD related cognitive function[1]. The composite included summary scores from five measures: (1) the MMSE [2], a commonly used test of global cognitive functioning; (2) the Logical Memory Delayed Recall (LMDR) [3], a story recall test following a 30 minute delay from initial presentation; (3) Digit-Symbol Coding (DSC)[4], a symbol substitution test that requires participants to fill in small blank squares that each have 1 of 9 numbers printed above, with shapes based on a key linking shapes with numbers; (4) the Free and Cued Selective Reminding Test, a multimodal associative memory test in which learning is enhanced by providing a visual and semantic category cue, in which participants are respond with free recall or cued with the semantic category, and the sum of free+total recall items is taken (FCSRT96)[5]; and (5) the Category Fluency test (CAT), in which participants generate words in 60 second belonging to a semantic category, with three categories administered (animals, vegetables, fruits).

The Clinical Dementia Rating scale (CDR) [6], a semi-structured interview with the participant and their caregiver was used to assess the severity of cognitive symptoms of dementia. The test was completed by experienced research staff, and scored based on the CDR Global Score (CDR-GS) and the CDR Sum of Boxes (CDR-SoB) rating scales.

Modifications to clinical assessments to adapt to a telemedicine setting are described in detail below.

**Supplementary table S1: Adjustments made to clinical assessments to adjust to a remote setting**

| Study procedure | Adaptations for remote assessment |
|---|---|
| MMSE | For virtual visits the MMSE was administered according to official administration material, with the following minor adaptations to the virtual setting:<br>● The spatial orientation questions include the questions "What room are you in?", referring to the specific room in the house the participant is within the house "e.g. living room".<br>● The object naming question requires showing the object either through the video camera, or presenting slides to the participant through Zoom's screen share function.<br>● The written command needs to be presented through the screenshared presentation material.<br>● The participant has the option to write the sentence through Zoom chat (or on paper that will then be shown through the camera).<br>● The intersecting pentagons to copy need to be screenshared, and the participant needs to show their drawing to the computer's camera. The participant is reminded to not make their drawing too small. |
| LMDR (Logical Memory Delayed Recall) - immediate recall | none |
| DSC (Digit Symbol Coding test) | The original DSC requires a piece of paper to be filled out by the participant, and was thus not feasible for remote administration in its original form. A digital adoption of the test was administered.<br><br>The participant was presented via screen share with a screen similar to the paper layout of original DSC: this will have the key (correct numbers for each symbol) in the top, and a number of rows below to assess digit coding. The digital adaptation differs in the way that each field in a row contains the symbol from the key, rather than the number, and there are no blank fields. The participant pairs up the symbols and the numbers, by speaking out the number associated with a given symbol in the row (as opposed to writing the symbol associated with the number). The main difference from the original test is swapping the role of the numbers of symbols; this has been done to enable verbal responses (where naming a number is feasible, but describing the figure each time is not).<br><br>The researcher administered the test using a set of powerpoint slides containing the digital adaption of the test. A coloured square highlights one of the fields in a row to show the participant what figure that should give the corresponding number to; once they answered, the researcher pressed a key and the coloured square moved to the next field. The researcher had a paper copy of the test where participant responses were written down. |
| FCSRT96 (Free and Cued Selective Reminding Test) | The FCSRT96 consists of presenting a range of cards with items on them. To adapt to the remote format, the clinician was provided with a powerpoint presentation where each slide corresponds to a card. Each slide contained four pictures were labelled with what corner of the screen they appear in (e.g. "top left"). During administration of the test (study phase), these slides were screen shared with the participant through the Video Platform. |
| CAT (Semantic Category Fluency Test) – animals, vegetables, fruits | None |
| Letter Fluency Test – F, A, S | None |
| Digit Span Backwards | None |
| LMDR - delayed recall | None |
| Clinical Dementia Rating Scale (CDR) - caregiver interview | A number of the questions on the CDR (in the sections "Orientation", "Patients outer activities") relate to activities of daily living that are expected to be impacted by quarantine that's been in place during the SARS-CoV2 pandemic. This were mitigated in the following ways: |

| | |
|---|---|
| Clinical Dementia Rating Scale (CDR) - participant interview | 1) The Rater was made aware that this is a potential issue, and during what parts of the interview extra attention should be paid to this. These sections are:<br>a) Orientation: question 6 and 7.<br>b) Patients outer activities: question: 1, 6, 7, 8.<br>2) The informant/participant was made aware of this at the start of the interview that questions should be answered in the context "in the hypothetical situation there were not any restrictions due to the pandemic".<br>3) The Rater could slightly rephrase questions to ask about the same matter, but in the context of the quarantine.<br>4) Scores were based on the Rater's judgement in the CDR test. The Rater could thus use common sense to not rely on an answer for scoring a section, if the answer wasn't valid given the quarantine. |

#### 3. Schedule of ASRT remote assessments

ASRTs were administered at the start of every remote assessment. Due to high burden in the original remote monitoring design, as fed back by participants, the assessment schedule was changed part-way through the study, favouring the use of shorter stories to reduce participant burden. Additional assessments which followed the initial ASRTs, not reported here, were reduced. Simultaneously, the number of days in which ASRT performance was assessed was increased from seven to eight days. Test schedule is documented below:

**Supplementary table S2: Schedule of self assessments.**

* Tasks with delayed recall after distraction task (category or verbal fluency). Each number-letter combination represents a different story stimulus. Further information on each story is provided in Skirrow et al. *submitted* [7]. Stories prefixed by "l" are long stories, and those prefixed by "s" are short stories.

| **Testing schedule** | **Remote Assessment Day** | | | | | | | |
|---|---|---|---|---|---|---|---|---|
| | **1** | **2** | **3** | **4** | **5** | **6** | **7** | **8** |
| Schedule 1: ASRT stories administered (prior to February 27th) | l1<br>l2<br>l3 | l4<br>l5<br>l6 | l7<br>l8<br>l9 | l10*<br>l11*<br>l12* | l13*<br>l14*<br>l15* | s1<br>s2<br>s3 | s4<br>s5<br>s6 | - |
| Schedule 2: ASRTs administered (after February 27th). | l1<br>l2<br>l3 | s1<br>s2<br>s3 | s4<br>s5<br>s6 | s7*<br>s8*<br>s9* | s10*<br>s11*<br>s12* | l10*<br>l11*<br>l12* | l13*<br>l14*<br>l15* | l4<br>l5<br>l6 |

#### 4. Prescreening simulation model

For simulation analysis we used population estimates for MCI prevalence documented in prior meta-analysis [8], broken down by 5-year age bands from age 65 to 85+. We aimed to simulate improvement in primary care referral based on the AI model compared with standard of care assessments completed via physician judgement (sensitivity=0.50, specificity=0.66) and the MMSE (sensitivity=0.51, specificity=0.71), using previously published results[9], and sensitivity and specificity of the AI system (short stories, immediate recall, ASR transcription).

We simulated a sample of 1000 people, divided into 4 year buckets from age 65 to 85. Group sizes were allocated proportionally to their representation in the US population in July 2020, as reported by Statista (https://www.statista.com/statistics/241488/population-of-the-us-by-sex-and-age/) represented by that age group (n=321 aged 65-70, n=263 aged 70-75, n=179 aged 75-80, n=116 aged 80-85, n=120 aged 85+).

Modelling was first carried out in individual age groups. Total positive cases (totalP) were calculated as n x prevalence for each age group, and total negatives (totalN) as n – totalP.

Test performance of different clinical use cases was calculated as follows, with *a* denoting age group, and *b* denoting the prediction model (AI model or Clinical Standard).

Total positive cases: $totalP_{a,b} = n_a \times prevalence_{a,b}$

Total negative cases: $totalN_{a,b} = n_a - totalP_{a,b}$

True positives: $TP_{a,b} = totalP_{a,b} \times$ model $sensitivity_b$

False negatives: $FN_{a,b} = totalP_{a,b} - TP_{a,b}$

True negatives: $TN_{a,b} = totalN_{a,b} \times$ model $specificity_b$

False positives: $FP_{a,b} = totalN_{a,b} - TN_{a,b}$

Difference in referral accuracy with using the AI system (*AI*) vs. clinical standard (*CS* - physician judgement or MMSE) was computed as follows using the sum (∑) across ages for TP, FP. Differences between

Difference in correct referrals: $\vartriangle C=TP_{AI,\ a},\ -\ TP_{CS,\ a}$

Difference in incorrect referrals: $\vartriangle I=FP_{AI,\ a},\ -\ FP_{CS,\ a}$

Proportional increase in true referrals: $PI= \sum \vartriangle C/\sum TP_{CS}$

Proportional decrease in false referrals: $PD=\sum \vartriangle I/\sum TP_{CS}$

#### 5. Demographic characteristics of subgroups

**Supplementary table S3: Participant demographic and clinical characteristics in full sample (N=133).**

Demographic and clinical characteristics shown by research groups 1-4, and summary statistics for participants characterised by clinical diagnostic or biomarker profiles. MCI: Mild Cognitive Impairment; AD: Alzheimer's Dementia; CU: Cognitively Unimpaired, N, Number; SD, standard deviation. Group1: Amyloid positive MCI/Mild AD, Group2: Amyloid negative MCI/Mild AD, Group 3: Amyloid positive cognitively unimpaired; Group 4: Amyloid negative cognitively unimpaired; MMSE: Mini Mental State Exam; CDR-G: Clinical Dementia Rating Scale - Global Score.

| | **Subgroup analyses** | | | | | **Full sample analyses** | | | | | |
|---|---|---|---|---|---|---|---|---|---|---|---|
| | | | | | | **Clinical group** | | | **Biomarker group** | | |
| | **Group 1 (N=31)** | **Group 2 (N=31)** | **Group 3 (N=36)** | **Group 4 (N=35)** | **p-value** | **CU (N=71)** | **MCI/mild AD (N=62)** | **p-value** | **Amyloid negative (N=66)** | **Amyloid positive (N=67)** | **p-value** |
| Amyloid positive/ Amyloid negative N | Positive | Negative | Positive | Negative | - | 36/35 | 31/31 | 0.94 | Negative | Positive | - |
| MCI/CU group or N | MCI | MCI | CU | CU | - | CU | MCI | - | 31/35 | 31/36 | 0.94 |
| Female/ Male (N) | 14/17 | 17/14 | 21/15 | 21/14 | 0.63 | 42/29 | 31/31 | 0.29 | 38/28 | 35/32 | 0.54 |
| Years in education, Mean (SD) | 14.83 (3.23) | 15.10 (3.13) | 15.08 (3.88) | 15.60 (3.34) | 0.83 | 15.34 (3.61) | 14.97 (3.16) | 0.46 | 15.38 (3.23) | 14.97 (3.57) | 0.46 |
| Age, Mean (SD) | 71.48 (5.30) | 67.77 (7.87) | 70.58 (4.10) | 69.914 (4.10) | 0.18 | 70.25 (4.09) | 69.63 (6.91) | 0.92 | 68.91 (6.21) | 71.00 (4.68) | 0.051 |
| MMSE, Mean (SD) | 27.20$^{A}$ (1.88) | 27.43$^{B}$ (2.01) | 29.19$^{C}$ (1.09) | 28.77$^{D}$ (1.07) | <0.001 AC, AD, BC, BD | 28.99 (1.10) | 27.32 (1.94) | <0.001 | 28.14 (1.71) | 28.29 (1.80) | 0.36 |
| CDR-G, mean (SD) | 0.53$^{A}$ (0.13) | 0.52$^{B}$ (0.16) | 0.07$^{C}$ (0.18) | 0.10$^{D}$ (0.21) | <0.001 AC, AD, BC, BD | 0.09 (0.19) | 0.53 (0.14) | <0.001 | 0.30 (0.28) | 0.28 (0.28) | 0.80 |

**Supplementary table S4: Participant demographic and clinical characteristics in the short story training subsample (N=110).**

Demographic and clinical characteristics shown by research groups 1-4, and summary statistics for participants characterised by clinical diagnostic or biomarker profiles. MCI: Mild Cognitive Impairment; AD: Alzheimer's Dementia; CU: Cognitively Unimpaired, N, Number; SD, standard deviation. Group1: Amyloid positive MCI/Mild AD, Group2: Amyloid negative MCI/Mild AD, Group 3: Amyloid positive cognitively unimpaired; Group 4: Amyloid negative cognitively unimpaired; MMSE: Mini Mental State Exam; CDR-G: Clinical Dementia Rating Scale - Global Score.

| | **Subgroup analyses** | | | | | **Full sample analyses** | | | | | |
|---|---|---|---|---|---|---|---|---|---|---|---|
| | | | | | | **Clinical group** | | | **Biomarker group** | | |
| | **Group 1 N=21** | **Group 2 N=26** | **Group 3 N=32** | **Group 4 N=31** | **p-value** | **CU N=63** | **MCI/mild AD N=47** | **p-value** | **Amyloid negative N=57** | **Amyloid positive N=53** | **p-value** |
| Amyloid positive/ Amyloid negative N | Positive | Negative | Positive | Negative | - | 32/31 | 21/26 | 0.53 | Negative | Positive | - |
| MCI/CU group or N | MCI | MCI | CU | CU | - | CU | MCI | - | 26/31 | 21/32 | 0.53 |
| Female/ Male (N) | 6/15 | 15/11 | 19/13 | 18/13 | 0.11 | 37/26 | 21/26 | 0.14 | 33/24 | 25/28 | 0.26 |
| Years in education, Mean (SD) | 15.10 (3.39) | 15.24 (2.86) | 15.06 (3.82) | 15.52 (3.35) | 0.98 | 15.29 (3.57) | 15.17 (3.08) | 0.78 | 15.39 (3.11) | 15.08 (3.62) | 0.78 |
| Age, Mean (SD) | 70.95 (5.97) | 67.19 (8.11) | 70.53 (4.27) | 69.68 (3.72) | 0.39 | 70.11 (4.00) | 68.87 (7.40) | 0.77 | 68.54 (6.19) | 70.70 (4.96) | 0.11 |
| MMSE, Mean (SD) | 27.29[A] (1.65) | 27.35[B] (2.04) | 29.22[C] (1.07) | 28.74[D] (1.12) | <0.001 AC, AD, BC | 28.98 (1.11) | 27.32 (1.85) | <0.001 | 28.11 (1.74) | 28.45 (1.62) | 0.21 |
| CDR-G, mean (SD) | 0.52[A] (0.11) | 0.50[B] (0.14) | 0.08[C] (0.19) | 0.10[D] (0.20) | <0.001 AC, AD, BC, BD | 0.09 (0.19) | 0.51 (0.13) | <0.001 | 0.29 (0.27) | 0.26 (0.27) | 0.6 |

**Supplementary table S5: Participant demographic and clinical characteristics in the long story training subsample (N=104).**

Demographic and clinical characteristics shown by research groups 1-4, and summary statistics for participants characterised by clinical diagnostic or biomarker profiles. MCI: Mild Cognitive Impairment; AD: Alzheimer's Dementia; CU: Cognitively Unimpaired, N, Number; SD, standard deviation. Group1: Amyloid positive MCI/Mild AD, Group2: Amyloid negative MCI/Mild AD, Group 3: Amyloid positive cognitively unimpaired; Group 4: Amyloid negative cognitively unimpaired; MMSE: Mini Mental State Exam; CDR-G: Clinical Dementia Rating Scale - Global Score.

| | **Subgroup analyses** | | | | | **Full sample analyses** | | | | | |
|---|---|---|---|---|---|---|---|---|---|---|---|
| | | | | | | **Clinical group** | | | **Biomarker group** | | |
| | **Group 1 (N=18)** | **Group 2 (N=26)** | **Group 3 (N=31)** | **Group 4 (N=29)** | **p-value** | **CU (N=60)** | **MCI/mild AD (N=44)** | **p-value** | **Amyloid negative (N=55)** | **Amyloid positive (N=49)** | **p-value** |
| Amyloid positive/ Amyloid negative N | Positive | Negative | Positive | Negative | - | 31/29 | 18/26 | 0.28 | Negative | Positive | - |
| MCI/CU group or N | MCI | MCI | CU | CU | - | CU | MCI | - | 26/29 | 18/31 | 0.28 |
| Female/ Male (N) | 7/11 | 15/11 | 19/12 | 18/11 | 0.40 | 37/23 | 22/22 | 0.24 | 33/22 | 26/23 | 0.48 |
| Years in education, Mean (SD) | 14.56 (3.29) | 15.16 (2.95) | 15.00 (3.93) | 15.28 (3.22) | 0.91 | 15.13 (3.58) | 14.91 (3.08) | 0.66 | 15.22 (3.07) | 14.84 (3.68) | 0.66 |
| Age, Mean (SD) | 70.94 (6.25) | 67.00 (8.02) | 70.39 (4.04) | 69.83 (3.61) | 0.35 | 70.12 (3.81) | 68.61 (7.53) | 0.64 | 68.49 (6.21) | 70.59 (4.91) | 0.12 |
| MMSE, Mean (SD) | 27.17$^{A}$ (1.51) | 27.46$^{B}$ (2.04) | 29.32$^{C}$ (0.91) | 28.72$^{D}$ (1.13) | <0.001 AC, AD, BC | 29.03 (1.06) | 27.34 (1.83) | <0.001 | 28.13 (1.73) | 28.53 (1.56) | 0.17 |
| CDR-G, mean (SD) | 0.53$^{A}$ (0.12) | 0.50$^{B}$ (0.14) | 0.08$^{C}$ (0.19) | 0.09$^{D}$ (0.20) | <0.001 AC, AD, BC, BD | 0.09 (0.19) | 0.51 (0.13) | <0.001 | 0.29 (0.27) | 0.25 (0.27) | 0.47 |

**Supplementary Figure S1: ROC curves for the AI system and baselines (short ASRTs, immediate recall)**

AUCs for the classifiers predicting: (A) amyloid, (B) mild cognitive impairment (MCI)/mild AD in the full sample. Subsample comparisons of classifier performance predicting (C) amyloid within the MCI; and (D) amyloid in the cognitively unimpaired (CU) sample. The table below each figure provides sensitivity and specificity at Youden's index, and Cohen's kappa measures. The reference test was biomarker confirmation on PET or CSF for A, C and D. Reference test was clinical diagnosis MMSE inclusion criteria for B. The demographic baseline includes age, sex and education level. ASR: automatic speech recognition - automatically transcribed; manual: manual transcription; AI: artificial intelligence, ASRT: automatic story recall test; PACC5: preclinical Alzheimer's cognitive composite with semantic processing; ROC: receiver operator characteristic (curve). AUC: area under the curve.

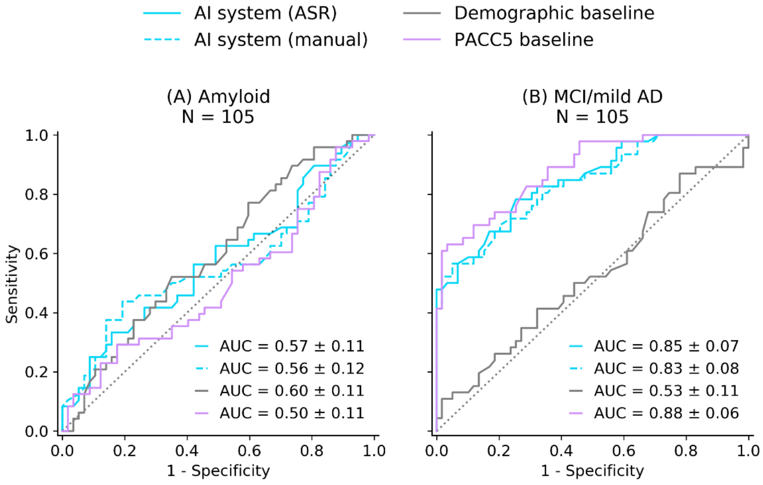


| | Sn | Sp | Cohen κ |
|---|---|---|---|
| AI (ASR) | 0.33 | 0.84 | 0.16 |
| AI (Manual) | 0.44 | 0.81 | 0.23 |
| Demographic | 0.77 | 0.40 | 0.13 |
| PACC5 | 0.29 | 0.82 | 0.10 |

| | Sn | Sp | Cohen κ |
|---|---|---|---|
| AI (ASR) | 0.78 | 0.75 | 0.50 |
| AI (Manual) | 0.57 | 0.95 | 0.51 |
| Demographic | 0.11 | 0.98 | 0.08 |
| PACC5 | 0.63 | 0.97 | 0.60 |

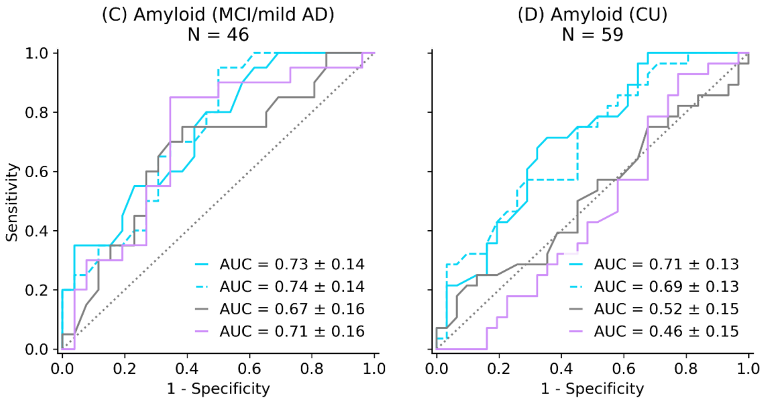


| | Sn | Sp | Cohen κ |
|---|---|---|---|
| AI (ASR) | 0.80 | 0.54 | 0.32 |
| AI (Manual) | 0.95 | 0.50 | 0.33 |
| Demographic | 0.75 | 0.62 | 0.31 |
| PACC5 | 0.85 | 0.65 | 0.44 |

| | Sn | Sp | Cohen κ |
|---|---|---|---|
| AI (ASR) | 0.71 | 0.65 | 0.36 |
| AI (Manual) | 0.75 | 0.55 | 0.26 |
| Demographic | 0.25 | 0.87 | 0.09 |
| PACC5 | 0.93 | 0.23 | 0.11 |

**Supplementary Figure S2: ROC curves for the AI system and baselines (short ASRTs, delayed recall)**

AUCs for the classifiers predicting: (A) amyloid, (B) mild cognitive impairment (MCI)/mild AD in the full sample. Subsample comparisons of classifier performance predicting (C) amyloid within the MCI; and (D) amyloid in the cognitively unimpaired (CU) sample. The table below each figure provides sensitivity and specificity at Youden's index, and Cohen's kappa measures. The reference test was biomarker confirmation on PET or CSF for A, C and D. Reference test was clinical diagnosis MMSE inclusion criteria for B. The demographic baseline includes age, sex and education level. ASR: automatic speech recognition - automatically transcribed; manual: manual transcription; AI: artificial intelligence, ASRT: automatic story recall test; PACC5: preclinical Alzheimer's cognitive composite with semantic processing; ROC: receiver operator characteristic (curve). AUC: area under the curve.

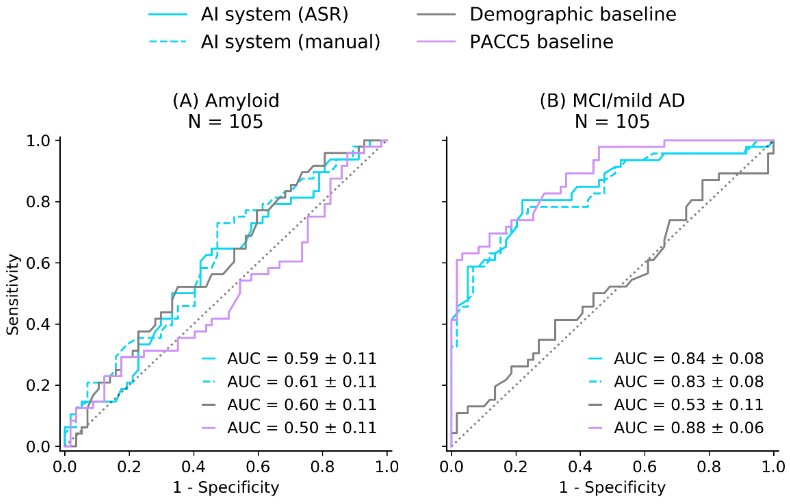


| | Sn | Sp | Cohen κ |
|---|---|---|---|
| AI (ASR) | 0.65 | 0.54 | 0.17 |
| AI (Manual) | 0.73 | 0.53 | 0.23 |
| Demographic | 0.77 | 0.40 | 0.13 |
| PACC5 | 0.29 | 0.82 | 0.10 |

| | Sn | Sp | Cohen κ |
|---|---|---|---|
| AI (ASR) | 0.80 | 0.78 | 0.54 |
| AI (Manual) | 0.78 | 0.76 | 0.52 |
| Demographic | 0.11 | 0.98 | 0.08 |
| PACC5 | 0.63 | 0.97 | 0.60 |

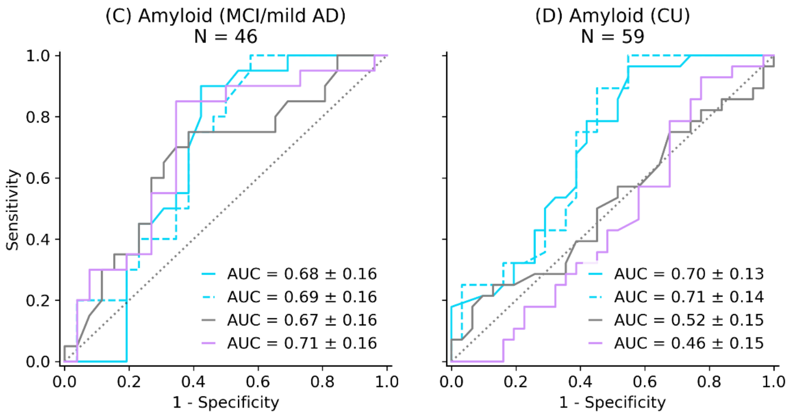


| | Sn | Sp | Cohen κ |
|---|---|---|---|
| AI (ASR) | 0.90 | 0.58 | 0.41 |
| AI (Manual) | 1.00 | 0.42 | 0.35 |
| Demographic | 0.75 | 0.62 | 0.31 |
| PACC5 | 0.85 | 0.65 | 0.44 |

| | Sn | Sp | Cohen κ |
|---|---|---|---|
| AI (ASR) | 0.96 | 0.45 | 0.37 |
| AI (Manual) | 1.00 | 0.45 | 0.37 |
| Demographic | 0.25 | 0.87 | 0.09 |
| PACC5 | 0.93 | 0.23 | 0.11 |

**Supplementary Figure S3: ROC curves for the AI system and baselines (long ASRTs, immediate recall)**

AUCs for the classifiers predicting: (A) amyloid, (B) mild cognitive impairment (MCI)/mild AD in the full sample. Subsample comparisons of classifier performance predicting (C) amyloid within the MCI; and (D) amyloid in the cognitively unimpaired (CU) sample. The table below each figure provides sensitivity and specificity at Youden's index, and Cohen's kappa measures. The reference test was biomarker confirmation on PET or CSF for A, C and D. Reference test was clinical diagnosis MMSE inclusion criteria for B. The demographic baseline includes age, sex and education level. ASR: automatic speech recognition - automatically transcribed; manual: manual transcription; AI: artificial intelligence, ASRT: automatic story recall test; PACC5: preclinical Alzheimer's cognitive composite with semantic processing; ROC: receiver operator characteristic (curve). AUC: area under the curve.

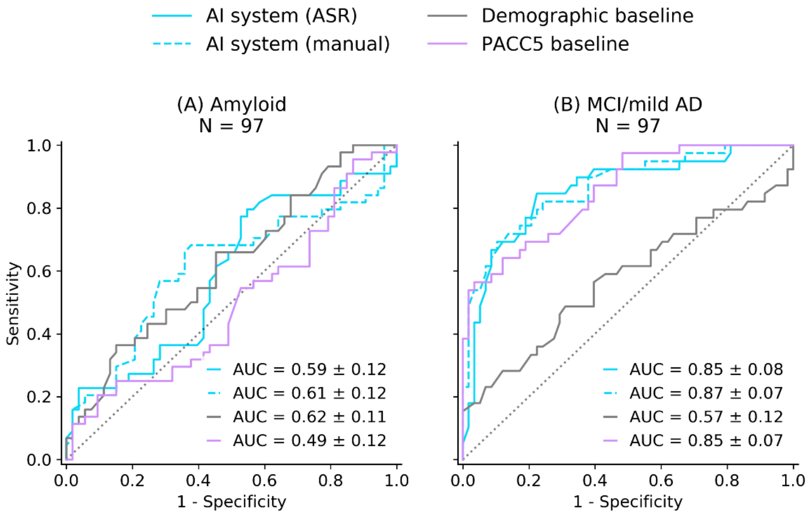


| | Sn | Sp | Cohen κ |
|---|---|---|---|
| AI (ASR) | 0.80 | 0.45 | 0.22 |
| AI (Manual) | 0.68 | 0.62 | 0.30 |
| Demographic | 0.36 | 0.85 | 0.20 |
| PACC5 | 0.20 | 0.91 | 0.09 |

| | Sn | Sp | Cohen κ |
|---|---|---|---|
| AI (ASR) | 0.85 | 0.78 | 0.56 |
| AI (Manual) | 0.69 | 0.90 | 0.58 |
| Demographic | 0.49 | 0.69 | 0.17 |
| PACC5 | 0.56 | 0.97 | 0.54 |

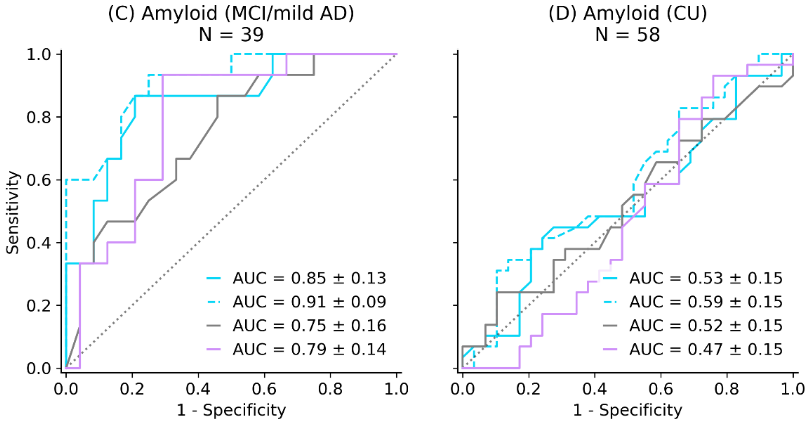


| | Sn | Sp | Cohen κ |
|---|---|---|---|
| AI (ASR) | 0.87 | 0.79 | 0.58 |
| AI (Manual) | 0.93 | 0.75 | 0.59 |
| Demographic | 0.87 | 0.54 | 0.31 |
| PACC5 | 0.93 | 0.71 | 0.43 |

| | Sn | Sp | Cohen κ |
|---|---|---|---|
| AI (ASR) | 0.45 | 0.72 | 0.17 |
| AI (Manual) | 0.31 | 0.90 | 0.17 |
| Demographic | 0.24 | 0.90 | 0.10 |
| PACC5 | 0.93 | 0.24 | 0.14 |

#### Supplementary Figure S4: ROC curves for the AI system and baselines (long ASRTs, delayed recall)

AUCs for the classifiers predicting: (A) amyloid, (B) mild cognitive impairment (MCI)/mild AD in the full sample. Subsample comparisons of classifier performance predicting (C) amyloid within the MCI; and (D) amyloid in the cognitively unimpaired (CU) sample. The table below each figure provides sensitivity and specificity at Youden's index, and Cohen's kappa measures. The reference test was biomarker confirmation on PET or CSF for A, C and D. Reference test was clinical diagnosis MMSE inclusion criteria for B. The demographic baseline includes age, sex and education level. ASR: automatic speech recognition - automatically transcribed; manual: manual transcription; AI: artificial intelligence, ASRT: automatic story recall test; PACC5: preclinical Alzheimer's cognitive composite with semantic processing; ROC: receiver operator characteristic (curve). AUC=area under the curve.

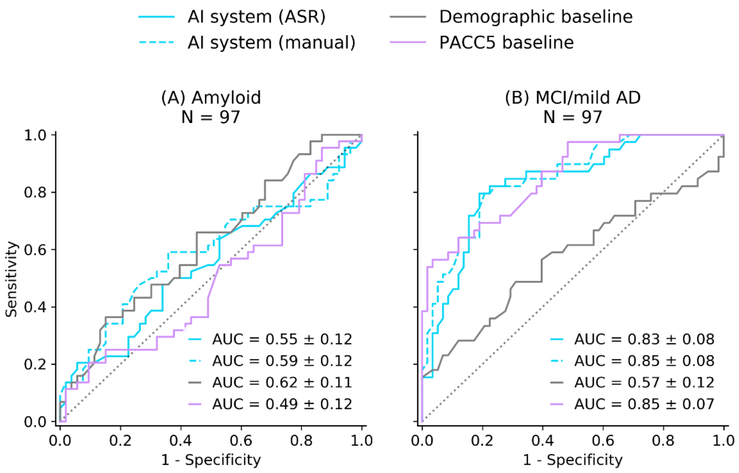


| | Sn | Sp | Cohen κ |
|---|---|---|---|
| AI (ASR) | 0.20 | 0.94 | 0.13 |
| AI (Manual) | 0.59 | 0.64 | 0.21 |
| Demographic | 0.36 | 0.85 | 0.20 |
| PACC5 | 0.20 | 0.91 | 0.09 |

| | Sn | Sp | Cohen κ |
|---|---|---|---|
| AI (ASR) | 0.79 | 0.81 | 0.57 |
| AI (Manual) | 0.79 | 0.79 | 0.55 |
| Demographic | 0.49 | 0.69 | 0.17 |
| PACC5 | 0.56 | 0.97 | 0.54 |

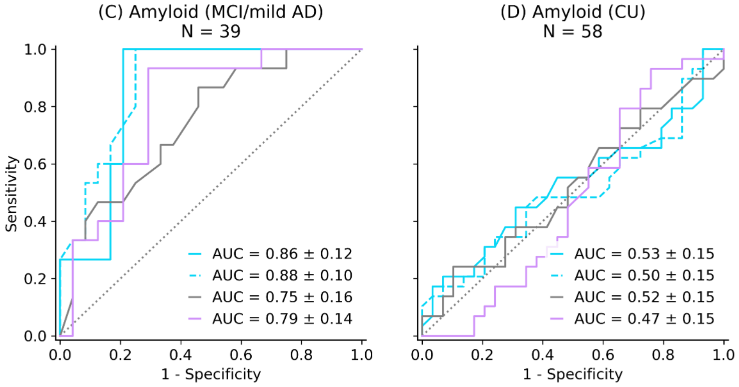


| | Sn | Sp | Cohen κ |
|---|---|---|---|
| AI (ASR) | 1.00 | 0.79 | 0.63 |
| AI (Manual) | 1.00 | 0.75 | 0.59 |
| Demographic | 0.87 | 0.54 | 0.31 |
| PACC5 | 0.93 | 0.71 | 0.43 |

| | Sn | Sp | Cohen κ |
|---|---|---|---|
| AI (ASR) | 0.17 | 0.97 | 0.10 |
| AI (Manual) | 0.48 | 0.62 | 0.10 |
| Demographic | 0.24 | 0.90 | 0.10 |
| PACC5 | 0.93 | 0.24 | 0.14 |

2 Folstein MF, Folstein SE, McHugh PR. "Mini-mental state" A practical method for grading the cognitive

state of patients for the clinician. *J Psychiatr Res* 1975; **12**: 189–98.

3 Wechsler D, Stone CP. Wechsler Memory Scale-Revised. San Antonio, TX: Psychological Corporation, 1987.

4 Wechsler D. WAIS-R Manual: Wechsler Adult Intelligence Scale-Revised. San Antonio, TX: Psychological Corporation, 1981.

5 Grober E, Ocepek-Welikson K, Teresi J. The Free and Cued Selective Reminding Test: Evidence of Psychometric Adequacy. *Psychol Sci Q* 2009; **51**: 266–82.

6 Morris JC. The Clinical Dementia Rating (CDR): current version and scoring rules. *Neurology* 1993; **43**: 2412–4.

7 Skirrow C, Meszaros M, Meepegama U, *et al.* Validation of a novel fully automated story recall task for repeated remote high-frequency administration. *MedrXiv* 2021.

8 Petersen RC, Lopez O, Armstrong MJ, *et al.* Practice guideline update summary: Mild cognitive impairment: Report of the Guideline Development, Dissemination, and Implementation Subcommittee of the American Academy of Neurology. *Neurology* 2018; **90**: 126–35.

9 Tong T, Thokala P, McMillan B, Ghosh R, Brazier J. Cost effectiveness of using cognitive screening tests for detecting dementia and mild cognitive impairment in primary care. *Int J Geriatr Psychiatry* 2017; **32**: 1392–400.
